# Conserved gene signatures shared among *MAPT* mutations reveal defects in calcium signaling

**DOI:** 10.1101/2022.06.10.22276260

**Authors:** Miguel A. Minaya, Sidhartha Mahali, Abhirami K. Iyer, Rita Martinez, John Budde, Sally Temple, Abdallah M. Eteleeb, Carlos Cruchaga, Oscar Harari, Celeste M. Karch

## Abstract

More than 50 mutations in the *MAPT* gene result in heterogeneous forms of frontotemporal lobar dementia with tau inclusions (FTLD-tau). However, early pathogenic events that lead to disease and the degree to which they are common across *MAPT* mutations remain poorly understood. The goal of this study is to determine whether there is a common molecular signature of FTLD-Tau. To do this, we analyzed genes differentially expressed in induced pluripotent stem cell (iPSC)–derived neurons that represent the three major categories of *MAPT* mutations: splicing (IVS10+16), exon 10 (p.P301L), and C-terminal (p.R406W) compared with isogenic controls. Here, we identified 275 genes that were commonly differentially expressed in *MAPT* IVS10+16, p.P301L, and p.R406W neurons. These genes were enriched in synaptic and endolysosomal pathways and neuronal development and were also altered in the presence of tau aggregation in a mouse model of tauopathy. The *MAPT* mutations commonly led to changes in genes and pathways sensitive to disruptions in calcium homeostasis. One of these genes, *CALB1*, plays a role in calcium dysregulation and is targeted by several FDA-approved drugs used to treat neurological symptoms. Finally, a subset of genes commonly differentially expressed across *MAPT* mutations were also dysregulated in brains from progressive supranuclear palsy patients, suggesting that molecular signatures relevant to genetic and sporadic forms of tauopathy are captured in a dish. The results from this study demonstrate that iPSC-derived neurons capture molecular processes that occur in human brains and can be used to pinpoint common molecular pathways involving synaptic and endolysosomal function and neuronal development, which may be regulated by disruptions in calcium homeostasis.

## Background

Frontotemporal lobal degeneration with tau inclusions (FTLD-tau) encompasses a heterogenous group of disorders characterized by frontal and temporal lobar atrophy, neuronal loss and gliosis, and the accumulation of neurofibrillary tangles (NFTs) (Bodea *et al*., 2016). In a subset of cases, FTLD-tau is caused by rare, dominantly inherited mutations in the *MAPT* gene (Pottier *et al*., 2016). Common genetic variation in the *MAPT* gene also contribute to sporadic forms of FTLD-Tau and (Steele *et al*., 2018; Kouri *et al*.; Ferrari *et al*.).

*MAPT* is alternatively spliced and developmentally regulated in the central nervous system, resulting in six canonical tau isoforms, which differ based on the inclusion of zero, one or two N-terminal inclusions (0N, 1N, 2N, respectively) and three or four repeats in the microtubule binding region (3R or 4R, respectively) (Neve *et al*., 1986; Lee, Goedert and Trojanowski, 2001). In adult brains, there is an equal balance of 3R and 4R tau isoforms. More than 50 mutations in the *MAPT* gene are reported to cause FTLD-tau (https://www.alzforum.org/mutations/mapt). These mutations fall into three major categories. First, located in the intronic region near the stem-loop domain, a subset of mutations alters *MAPT* splicing, inclusion of exon 10 (e.g. 3R<4R tau) or exclusion of exon 10 (e.g. 3R>4R tau). Second, missense mutations occurring within exon 10, such that the mutation is only present in *MAPT* isoforms containing exon 10 (e.g. 4R tau). Finally, missense mutations may occur outside of the microtubule binding domain, which leads to the production of mutant protein among all tau isoforms. We asked whether the heterogeneity in *MAPT* mutations drive distinct molecular mechanisms. To begin to address this question, we studied common *MAPT* mutations that fall into these three major categories: *MAPT* IVS10+16, p.P301L and p.R406W, respectively.

Human cellular models of the brain derived from iPSC have become an important tool for studying cellular markers that may initiate disease (Guo *et al*., 2017; Lagomarsino *et al*., 2021; Gonzalez *et al*., 2018; Iovino *et al*., 2015; Jiang *et al*., 2018; Nakamura *et al*., 2019; Silva *et al*., 2016; Wray, 2017; Bowles *et al*., 2021; Livesey, 2014; Karch *et al*., 2018). Here, we coupled human iPSC models with CRISPR/Cas9 genome editing technology to create a system that allows us to distinguish the molecular signatures associated with FTLD-tau and to begin to resolve the key molecular phenotypes of tauopathy. Together, our findings uncover key changes in calcium signaling shared across *MAPT* mutations.

## Materials and Methods

### Patient consent

Skin punches were performed following written informed consent from the donor. The informed consent was approved by the Washington University School of Medicine and University of California San Francisco Institutional Review Board and Ethics Committee (IRB 201104178, 201306108 and 10-03946). The consent allows for use of tissue by all parties, commercial and academic, for the purposes of research but not for use in human therapy.

### iPSC lines

Human iPSC used in this study (Supplemental Table 1; Supplemental Figure 1) have been previously described (Karch *et al*., 2019). Briefly, iPSC lines were generated using non-integrating Sendai virus carrying OCT3/4, SOX2, KLF4, and cMYC (Life Technologies) (Takahashi and Yamanaka, 2006; Ban *et al*., 2011). iPSC lines were characterized for the following parameters using standard methods (Takahashi and Yamanaka, 2006): pluripotency markers by immunocytochemistry (ICC) and quantitative PCR (qPCR), spontaneous or TriDiff differentiation into the three germ layers by ICC and qPCR, assessment of chromosomal abnormalities by karyotyping, and *MAPT* mutation status was confirmed by Sanger sequencing (Supplemental Figure 1). To determine the impact of the *MAPT* mutant allele on molecular phenotypes, we used CRISPR/Cas9-edited isogenic controls in which the mutant allele was reverted to the wild-type (WT) allele in each of the donor iPSC lines as previously described (GIH36.2; F11362.1; F0510.2; Supplemental Table 1; Supplemental Figure 1) (Karch *et al*., 2019). Resulting edited lines were characterized as described above in addition to on- and off-target sequencing (Supplemental Figure 1). All iPSC lines used in this study carry the *MAPT* H1/H1 common haplotype.

### Differentiation of iPSCs into cortical neurons

iPSCs were differentiated into cortical neurons using a two-step approach as previously described (Karch *et al*., 2019) (https://dx.doi.org/10.17504/protocols.io.p9kdr4w). iPSCs were plated at a density of 65,000 cells per well in neural induction media (StemCell Technologies) in a 96-well v-bottom plate to form neural aggregates and after 5 days, transferred into culture plates. The resulting neural rosettes were then isolated by enzymatic selection (Neural Rosette Selection Reagent; StemCell Technologies) and cultured as neural progenitor cells (NPCs). NPCs were differentiated in planar culture in neuronal maturation medium (neurobasal medium supplemented with B27, GDNF, BDNF, cAMP). Neurons typically arose within one week after plating, identified using immunocytochemistry for β-tubulin III (Tuj1). The cells continue to mature and were analyzed at 6 weeks.

### RNA extraction, sequencing and transcript quantification

iPSC-derived neurons were re-suspended in 200µL of 50:1 homogenization solution: 1-Thioglycerol solution. After addition of 200µL of Promega lysis buffer, the samples were transferred to the appropriate well of the Maxwell RSC cartridge. DNase solution was added to each cartridge. TapeStation 4200 System (Agilent Technologies) was used to perform quality control of the RNA concentration, purity, and degradation based on the estimated RNA integrity Number (RIN), and DV200 (Supplemental Table 1). Samples were sequenced by an Illumina HiSeq 4000 Systems Technology with a read length of 1x150 bp, and an average library size of 36.5 ± 12.2 million reads per sample

Identity-by-Descent (IDB) (Browning and Browning, 2010) and FastQC (Andrews *et al*., 2012) analyses were performed to confirm sample identity. STAR (v.2.6.0) (Dobin *et al*., 2012) was used to align the RNA sequences to the human reference genome: GRCh38.p13 (hg38). The quality of RNA alignment was evaluated using sequencing metrics such as read distribution, ribosomal content, and alignment quality in Picard (v.2.8.2). The average percentage of unique mapped reads in the BAM files was 80.3% ± 3.62, and the average percentage of total mapped reads to GRCh38.p13 was 90.1% ± 5.12 (Supplemental Table 1). IGV (Integrative Genomics Viewer) (Thorvaldsdottir, Robinson and Mesirov, 2013) was used with the reference Human Genome (hg38) to visualize mutation containing reads and their absence in samples edited using CRISPR/Cas9 protocols (isogenic controls).

Salmon (v. 0.11.3) (Patro *et al*., 2017) was used to quantify the expression of the genes annotated within the human reference genome used in this project (GRCh38.p13). Protein coding genes were selected for downstream analyses.

### Principal component and differential expression analyses

Principal component analyses (PCA) were performed based on 19,957 protein coding genes using regularized-logarithm transformation (rlog) counts. Differential gene expression was performed using the DESeq2 (v.1.22.2) R package (Love, Huber and Anders, 2014). PCA and differential gene expression analyses were performed independently for each set of *MAPT* mutations and isogenic controls. Each *MAPT* mutation and its isogenic control were considered independent cohorts due to their shared genetic background. As such, the relationship across the three *MAPT* mutation sets was evaluated using the *MetaVolcanoR* R package (v1.10.0) (Prada, Lima and Nakaya, 2021). The meta-analysis included those genes that were differentially expressed (p<0.05) in the same direction across the three cohorts (n= 275 genes). A meta-volcano plot summarizing the gene fold change of the *MAPT* IVS10+16, p.P301L, and p.R406W datasets was generated using a Random Effect Model (REM) estimation. PCA and Volcano plots were created for each comparison using the ggplot2 R package (v3.3.6) (Wickham, 2016).

### Pathway enrichment and network analyses

ToppGene (Chen *et al*., 2009) and Enrichr (Chen *et al*., 2013; Kuleshov *et al*., 2016; Xie *et al*., 2021) were used to identify pathways in which differentially expressed genes are enriched. Gene ontologies (GOs) related to molecular function, biological process and cellular component were selected based on two criteria: (i) p ≤ 0.05 and (ii) number of query genes associated with each GO > 1. Gene relationships including physical, predicted and genetic interactions, and gene networks including co-expression and co-localization were annotated using the geneMANIA prediction server (Warde-Farley *et al*., 2010).

### Mouse model of tauopathy

To evaluate whether the genes differentially expressed in iPSC-derived neurons were altered in animal models of tauopathy, we analyzed the gene expression in the Tau-P301L mouse model of tauopathy and non-transgenic controls (Ramsden *et al*., 2005). Transcriptomic data from mice was obtained from the Mouse Dementia Network (Matarin *et al*., 2015). Gene expression across the timepoints (2-, 4-, 8-, and 18-months old mice) was normalized to mice at two months of age and plotted. Differential gene expression at 18 months of age was analyzed by unpaired t-tests to assess significance.

### Drug target identification

To determine whether differentially expressed genes were associated with known to drugs, we interrogated: (i) the WEB-based Gene SeT AnaLysis Toolkit (Liao *et al*., 2019), (ii) the Drug-Gene Interaction Database (Freshour *et al*., 2021), and (iii) the DrugBank (Wishart *et al*., 2018).

### Calcium imaging

To measure calcium levels in iPSC-derived neurons, *MAPT* IVS10+16 mutation (GIH36.2) and isogenic controls (GIH36.2Δ1D01) were analyzed. NPCs were differentiated into cortical neurons as described above. After 22 days in culture, 2x10^5^ neurons of each genotype were seeded into poly-L-ornithine and laminin-coated 96-well plate. Ca^2+^ levels in the iPSC-derived neurons were then measured using the Invitrogen™ Fluo-4 Direct™ Calcium Assay Kit (catalog number: F10471) following manufacturer’s instructions. Briefly, at 36 days in culture, growth medium was replaced with 50μL per well Fluo-4 Direct^TM^ calcium assay buffer and 50μL per well of the 2x Fluo-4 Direct^TM^ calcium reagent loading solution. The 96-well plate was then incubated at 37°C for 60 minutes, after which fluorescence was measured using Synergy HTX multi-mode microplate reader from BioTek Instruments using instrument settings for excitation at 494nm and emission at 516nm. Negative controls included Fluo-4 Direct^TM^ calcium assay buffer plus reagent with no neurons and neurons without assay reagent. Fluo-4 staining in cells was imaged under a Nikon Eclipse 80i fluorescent microscope at 20x magnification. Cells were lysed in 1% Triton X-100 and total fluorescence intensities were measured as described above.

### PSP brains

To determine whether the differentially expressed genes in the iPSC-derived neurons capture molecular processes that occur in human brains with primary tauopathy, we analyzed gene expression in a publicly available dataset: the temporal cortex of 76 control and 82 PSP brains (syn6090813)(Allen *et al*., 2016). Differential gene expression analyses comparing controls and PSP brains was performed using a “Simple Model” that employs multi-variable linear regression analyses using normalized gene expression measures and corrected by gender, age-at-death, RNA integrity number (RIN), brain tissue source, and flowcell as covariates (Allen *et al*., 2016).

## Results

### MAPT mutations are sufficient to induce global transcriptomic changes in human neurons

The goal of this study was to identify common genes and pathways that are downstream of the *MAPT* mutation and candidate drivers of disease pathogenesis in FTLD-tau (Figure 1). To address this goal, we studied a series of *MAPT* mutations that represent three major categories: *MAPT* IVS10+16, p.P301L and p.R406W (Figure 1; Supplemental Table 1). We analyzed protein coding genes obtained from RNA-sequencing data generated from induced pluripotent stem cell (iPSC)–derived neurons carrying one of these three *MAPT* mutations together with isogenic controls (Figure 2A). PCAs illustrate that each of the *MAPT* mutations produced a robust effect on gene expression and were sufficient to induce global transcriptomic changes in iPSC-neurons: 81.66% principal component 1 (PC1) for *MAPT* IVS10+16; 79.33% PC1 for *MAPT* p.P301L; and 57.28% PC1 for *MAPT* p.R406W (Figure 2B, 2D and 2F). Given that the genomic background remains conserved within the pairs, we treated each pair as a cohort and performed differential expression analyses to determine the impact of the presence of each mutant allele (Figure 2C, 2E and 2G; Supplemental Tables 2 and 3). Together, these findings illustrate that FTD-causing *MAPT* mutations are sufficient to produce robust gene expression changes in neurons.

**Figure 1.**
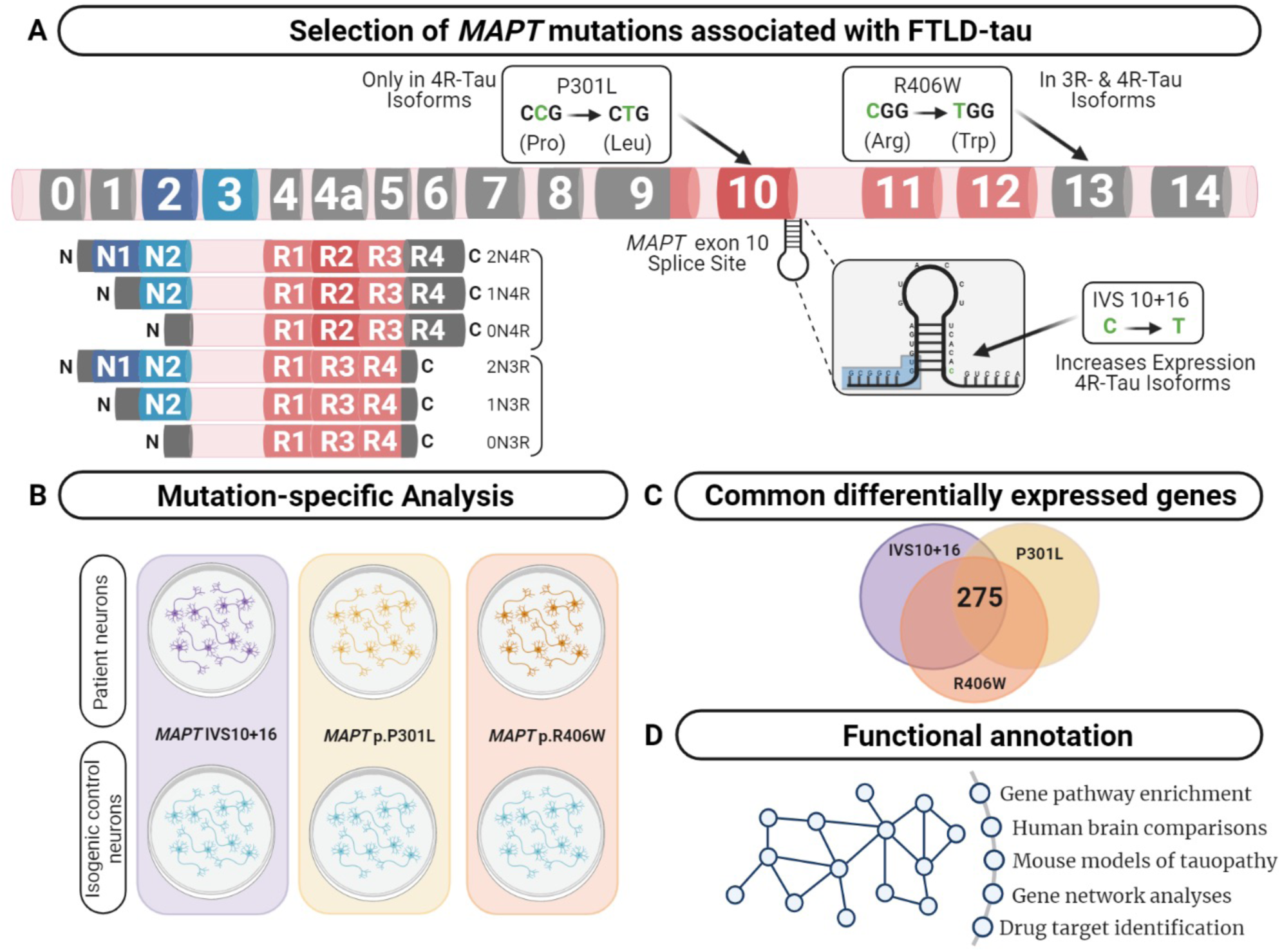
Integrative analysis to identify dysregulated pathways in FTD-Tau. (A) *MAPT* gene annotated with the location of the mutations used in this study. Lower left panel displays the 6 major isoforms expressed in the central nervous system. (B) Comparison of human iPSC- neurons carrying the *MAPT* mutation specific and isogenic controls served as a discovery cohort to identify genes dysregulated across the three mutations. (C) Overlap analysis between all mutants compared with control. Unadjusted p-values (p ≤ 0.05) revealed 275 commonly differentially expressed genes. (D) Functional annotation was performed using the commonly differentially expressed genes.

**Figure 2.**
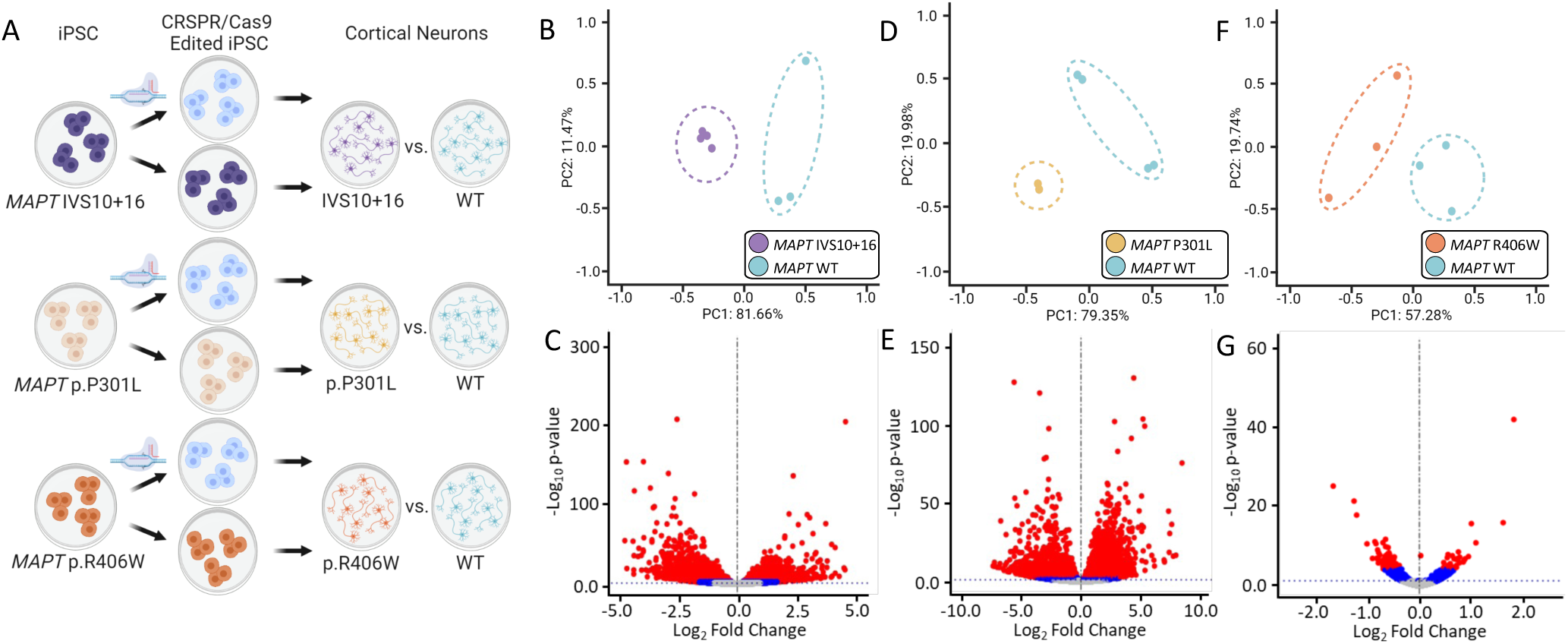
Global transcriptomic effects of *MAPT* IVS10+16, p.P301L, and p.R406W mutations. (A) Overview of iPSC differentiation into cortical neurons. (B – C) Principal component analyses and volcano plots obtained from *MAPT* IVS10+16 neurons compared to isogenic controls. (D – E) PCA and Volcano plots obtained from *MAPT* p.P301L neurons compared to isogenic controls. (F – G) PCA and Volcano plots obtained from *MAPT* p.R406W neurons compared to isogenic controls. PCA and Volcano plots were based on 19,957 protein-coding genes using regularized-logarithm transformation (rlog) counts. Volcano plots showing log_2_fold change between iPSC- derived neurons carrying *MAPT* mutations vs isogenic controls, and the –log_10_ p-value for each gene. Red and blue dots within volcano plots represent, respectively, genes differentially expressed under adjusted-BY (BY-FDR) and unadjusted p-values (p ≤ 0.05).

### MAPT mutations produce a shared gene expression signature in human neurons

Differential gene expression analyses within isogenic pairs illustrates that individual *MAPT* mutations produce global transcriptomic changes; thus, we sought to determine the extent of overlap in the differentially expressed genes among these three distinct classes of *MAPT* mutations (Figure 1C). We identified 275 differentially expressed genes across the three datasets. Of those, 159 were up-regulated and 116 down-regulated genes (p< 0.05; Figure 3A, Supplemental Table 4). To reduce the rate of false positives, we performed a meta-analysis (Bravata and Olkin, 2001)(Figure 3B). Pathway analyses revealed that the 159 up-regulated genes were enriched for (i) trans-synaptic signaling pathways such as negative regulation of canonical Wnt signaling; (ii) neuronal projection pathways such as forebrain development; and (iii) endolysosomal pathways such as endonuclease activity (Figure 3C). The 116 down-regulated genes were enriched for (i) trans-synaptic signaling pathways such as cAMP-mediated signaling; (ii) neuronal projection pathways associated with learning and cognition; and (iii) endolysosomal pathways such as phagosome maturation, regulation of autophagy, and vesicle transport along microtubules (Figure 3C). These findings point to a common set of genes and pathways that are disrupted downstream of the three distinct classes of *MAPT* mutations.

**Figure 3.**
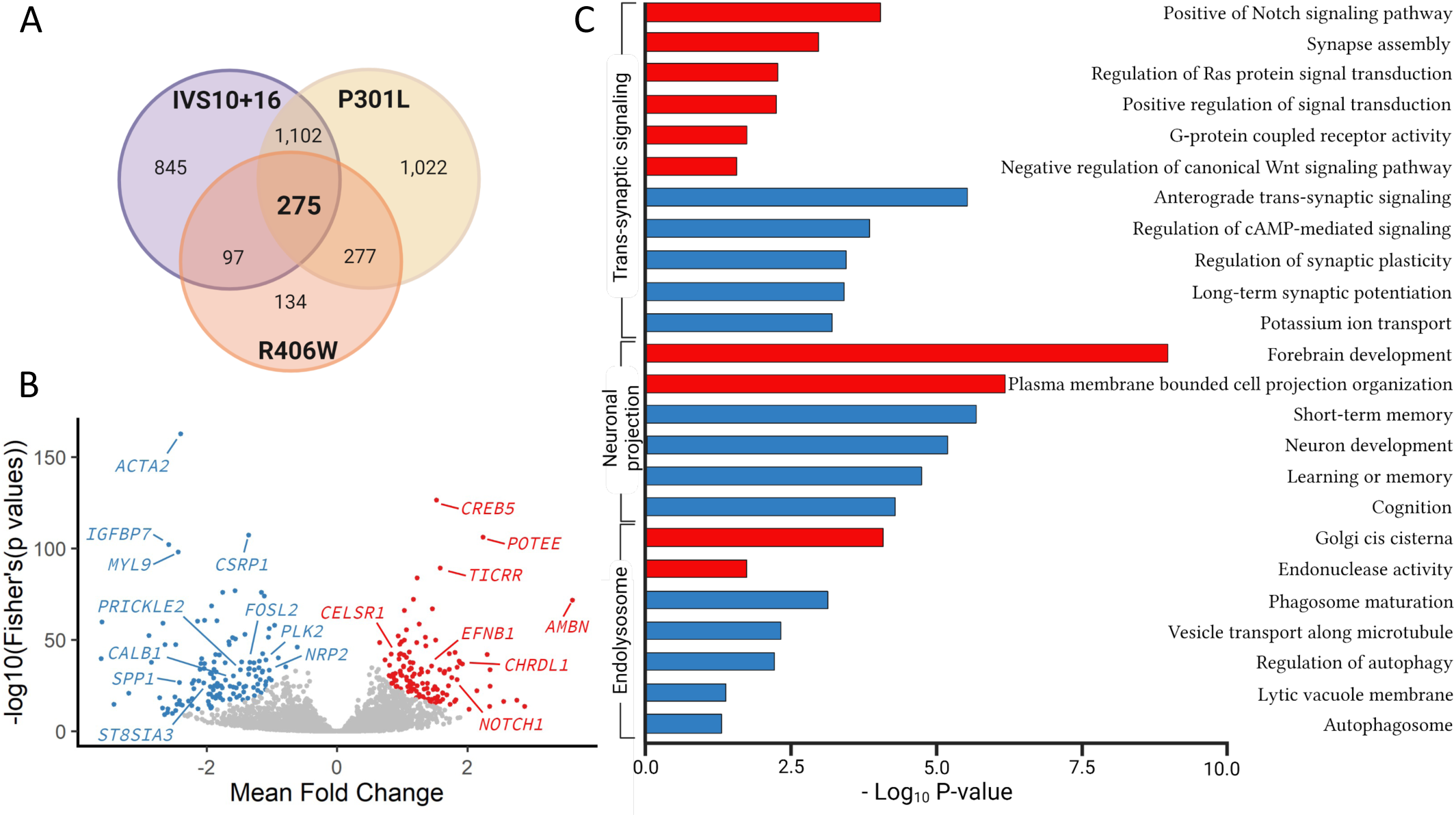
*MAPT* mutations result in common defects in synaptic signaling, neuronal projection, and endolysosomal function. (A) Venn diagram presenting the differentially expressed genes common among iPSC-neurons carrying *MAPT* IVS10+16, p.P301L and p.R406W mutations (p ≤ 0.05). (B) Meta-Volcano plot identifies the common gene expression changes of the *MAPT* IVS10+16, p.P301L and p.R406W. Red and blue dots represent up- and down- regulated genes, respectively. (C) Bar graph showing pathways enriched with the 159 up- and 116 down-regulated genes common among the three mutations. Red and blue bars represent, respectively, pathways associated with up- and down-regulated genes.

### Unique gene signatures

Beyond molecular signatures shared across the three *MAPT* mutations, we observed an imbalance in those genes shared between exon 10 mutations (*MAPT* IVS10+16 and p.P301L) and point mutations (*MAPT* p.P301L and p.R406W). We identified 499 genes shared between iPSC-neurons carrying the *MAPT* IVS 10+16 and p.P301L mutations (BY-FDR<0.05; Supplemental Figure 2 and Supplemental Table 5), while only 16 genes were shared between iPSC-neurons carrying the *MAPT* p.P301L and p.R406W mutations (Supplemental Figure 2 and Supplemental Table 6). Thus, the number of differentially expressed genes associated with mutations located around the alternatively spliced exon 10 was 31-fold higher than the number of dysregulated genes associated with the *MAPT* p.P301L and p.R406W mutations.

The 235 up-regulated genes found in the *MAPT* IVS10+16 and p.P301L neurons revealed pathways involved in the regulation of cell signaling: (i) receptor signaling via JAK-STAT pathway; (ii) Rap protein signal transduction; and (iii) ERBB signaling pathway (Supplemental Figure 2; Supplemental Table 7). Among the 264 down-regulated genes in the *MAPT* IVS10+16 and p.P301L neurons, a number of genes were enriched in pathways involved in endolysosomal function: (i) the coated vesicle membrane; (ii) secretory vesicles; (iii) lysosome; and (iv) vacuolar lumen (Supplemental Figure 2; Supplemental Table 7). The genes shared among the *MAPT* p.R406W and p.P301L neurons were enriched in pathways related to neurogenesis and synaptic function (Supplemental Figure 2; Supplemental Table 8).

### MAPT mutations lead to common genetic signatures that are associated with tau aggregation in mouse models of tauopathy

We sought to determine the extent to which the genes commonly differentially expressed across *MAPT* mutations were altered during disease course in the Tau-P301L mouse model of tauopathy. Here, we restricted our analyses to those genes that were commonly differentially expressed in the *MAPT* mutant neurons (BY-FDR ≤ 0.05; Figure 4A; Supplemental Table 9). We identified 11 genes differentially expressed in the same direction across the three *MAPT* mutations (Figure 4B-D). Using the Mouse Dementia Network (Matarin *et al*., 2015), we then analyzed transcriptomic data generated from the cortex of WT and Tau-P301L mice collected at 2, 4, 8, and 18-months (Figure 4E). Compared with WT littermates, transgenic Tau-P301L mice develop cognitive impairments from 4 months of age and form NFTs with increasing age (Ramsden *et al*., 2005). We found that 6 of the 11 genes (*Celsr1*, *Chrdl1*, *Calb1*, *Plk2*, *Prickle2*, *St8sia3*) were differentially expressed in the tauopathy mouse model at 18 months of age, when tau aggregation is most prominent (Figure 4F-K; Supplemental Figure 3). These 6 genes are highly related to one another and enriched in pathways related to neurodegenerative diseases such as neurogenesis, behavior, learning, memory, and glutamatergic synapse (Figure 4L; Supplemental Table 10). The Drug-Gene Interaction Database (Freshour *et al*., 2021) and DrugBank (Wishart *et al*., 2018) revealed that two genes, *PLK2* and *CALB1*, are known targets of FDA approved drugs including tramadol, ethosuximide, levodopa, nicotine and oxcarbazepine, which are currently utilized to treat neurological symptoms (Supplemental Table 11). Together, these findings illustrate that *MAPT* mutations are sufficient to induce molecular changes *in vitro* that are relevant to disease pathogenesis and identify therapeutic targets for which there are FDA approved drugs.

**Figure 4.**
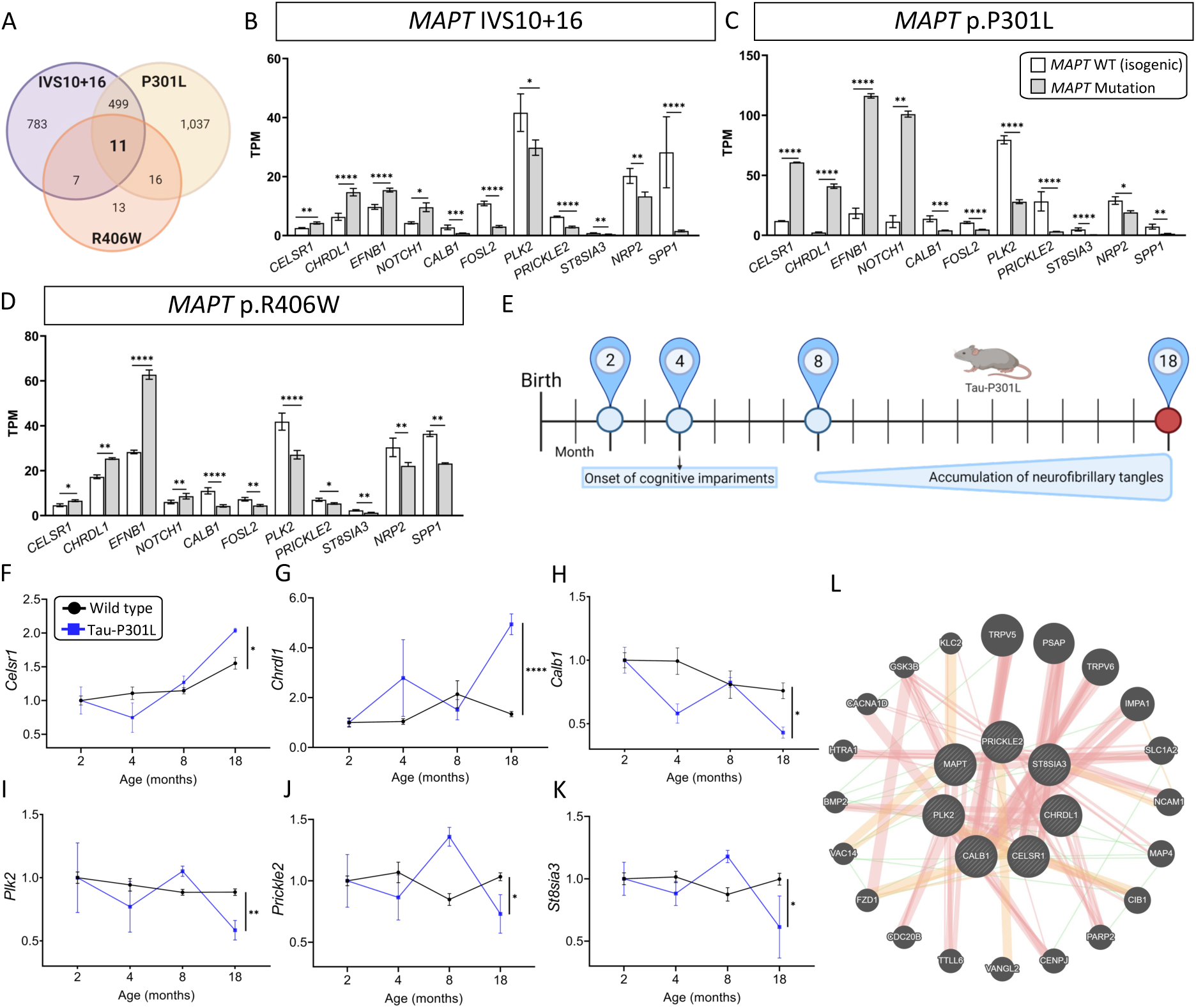
Genes altered in iPSC-neurons from *MAPT* mutation carriers are replicated in disease end-stage in the Tau-P301L tauopathy mouse model. (A) Venn Diagram presenting the differentially expressed genes overlapping between neurons carrying *MAPT-*IVS10+16, p.P301L and p.R406W mutations using adjusted BY-FDR ≤ 0.05. (B – D) Normalized TPM expression of 11 high confidence genes overlapped between the three datasets (*MAPT-* IVS10+16, p.P301L and p.R406W; BY-FDR ≤ 0.05). (E) The expression of the 11 genes observed in iPSC-neurons were evaluated in the mouse model of tauopathy (Tau-P301L vs wild type). Statistical comparisons reflect gene expression at disease end-stage (18 months old mice). (F – K) Expression of the 6 selected genes (2 up- and 4 down-regulated) in mice. Black circles, transgenic Tau-P301L mice; blue squares, non-transgenic control mice. Graphs show normalized gene expression relative to two months old mice. The genes presented were selected based on: (i) the expression of the genes in the human cell models and in mice followed the same direction, and (ii) the difference in the expression of each gene in 18 months old controls and transgenic Tau-P301L mice was significant. Statistical analyses (t-tests) were based on values normalized related to mice that were two months old. *p ≤ 0.05; **p ≤ 0.005; ****p ≤ 0.00005. (L) Relatedness of the 6 selected genes dysregulated in human and mice, including the *MAPT* gene. The plot demonstrates the strong physical (orange nodes: 80.3%) and genetic interactions (green nodes 4.28%) between the query genes and genes have been related to pathways such as neurogenesis, behavior, learning or memory, and glutamatergic synapse (Supplemental Table 8). The size of the gene nodes is proportional to the degree to which the genes are related. While query genes are presented as striped dark grey balls, other selected genes subjected to interaction are presented as dark grey balls without stripes. Orange, yellow, and green lines represent, respectively, physical (80.3%), predicted (9.48%), and genetic (4.28%) interactions within the GeneMANIA network.

### Gene dysregulation downstream of MAPT mutations impact calcium content

We next sought to understand the functional consequences of *MAPT* mutation driven gene changes. We focused on the six genes, *CELSR1, CHRDL1, CALB1, PLK2, PRICKLE2, ST8SIA3,* that were found to be commonly differentially expressed across *MAPT* mutations in iPSC-neurons and were altered with tau aggregation in the Tau-P301L mouse model. A common feature of calcium dysregulation emerged. In particular, the *CALB1* gene, which encodes the calbindin 1 protein and regulates Ca^2+^ entry into cells upon the stimulation of glutamate receptors (Noble *et al*., 2018), was significantly down-regulated in *MAPT* mutant iPSC-neurons (Figure 5A) and in 18-month old Tau-P301L mice (Figure 5B). Thus, we hypothesized that reduced *CALB1* expression leads to reduced intracellular calcium. To test this hypothesis, we measured calcium levels in iPSC-neurons from *MAPT* IVS10+16 and isogenic controls. We observed a significant reduction in calcium levels in *MAPT* IVS10+16 iPSC-neurons which was maintained in the total calcium pool (Figure 5C-D). Thus, we show that *MAPT* mutations are sufficient to disrupt calcium homeostasis in neurons, possibly through the reduction of *CALB1*.

**Figure 5.**
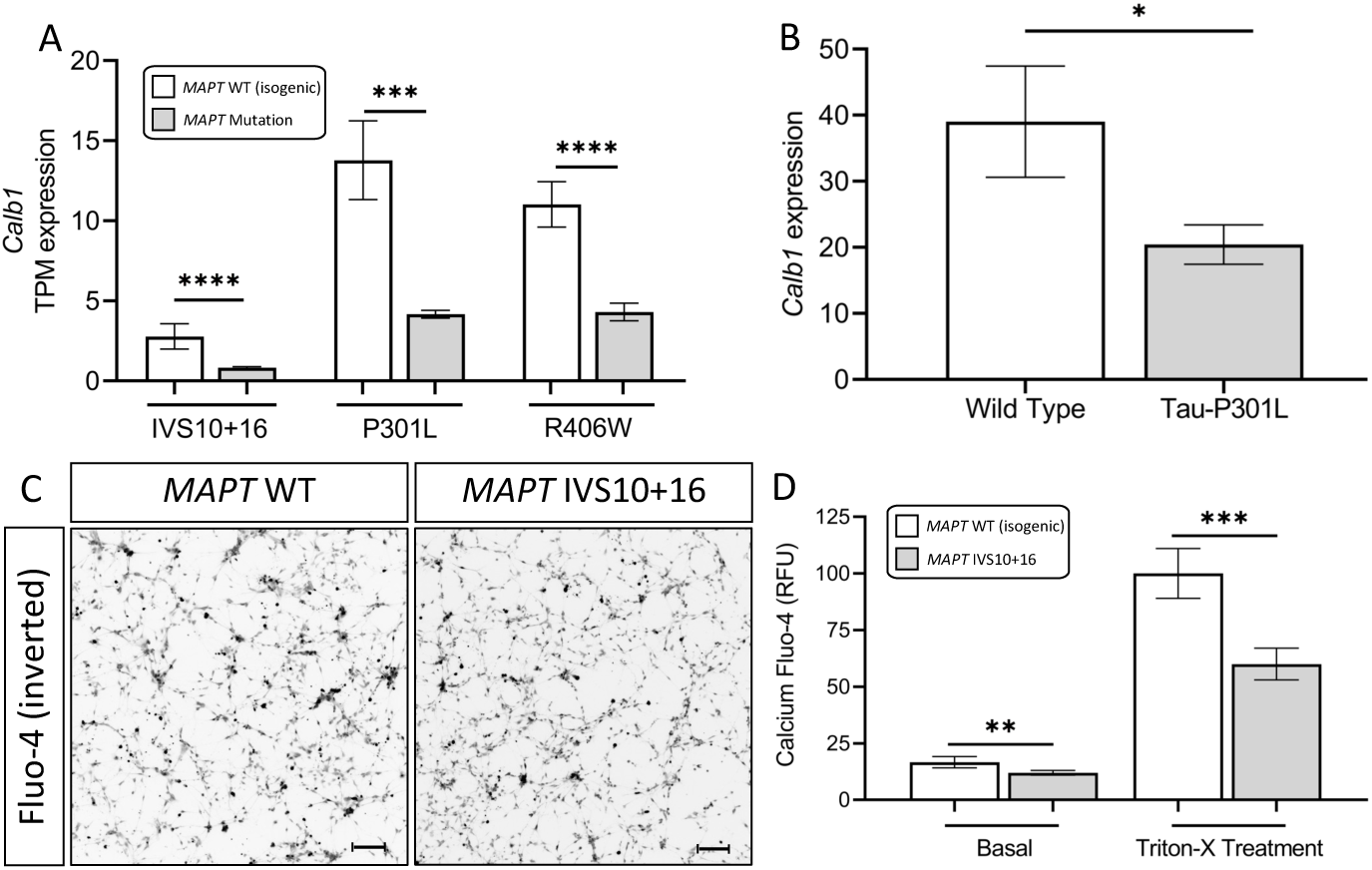
Calcium is dysregulated in *MAPT* neurons. (A) Normalized TPM expression of the *CALB1* gene in iPSC-neurons carrying the IVS10+16, p.P301L and p.R406W mutation. (B) Expression of the *Calb1* gene in 18-months-old WT vs Tau-P301L transgenic mouse model of tauopathy. (C-D) iPSC-neurons from *MAPT* IVS10+16 and isogenic control were cultured for 42 days and evaluated for intracellular calcium levels. (C) Inverted fluorescent imaging representing the calcium concentration in iPSC-neurons treated with Fluo-4 Direct™ Calcium Assay Kit. (D) Fluo4-Ca^2+^ binding dye relative fluorescence units (RFU) observed in iPSC-neurons carrying the IVS10+16 (grey bars) and isogenic controls (white bars) (*p ≤ 0.05; **p ≤ 0.005; ***p ≤ 0.0005; ****p ≤ 0.00005). Scale bar 20uM.

### Stem cell models capture disease-relevant gene signatures

Leveraging isogenic iPSC lines to understand the contribution of a single allele to downstream phenotypes is a powerful system. However, a limitation of this approach remains that the human neurons are cultured in a dish. To determine the extent to which the common gene signatures we observe in our cell lines are relevant to gene expression changes occurring in human brains with tauopathy, we compared the 275 differentially expressed genes (Figure 3A) to a transcriptomic dataset that includes control and PSP brains (Figure 6A)(Allen *et al*., 2016). We found that 43 of the 275 genes were also differentially expressed in the same direction in PSP brains (Figure 6A; Supplemental Table 12). Among the down-regulated genes (n=10), *VMP1* and *RABGAP1L* were associated with protein transport (GO:0015031). Network analyses showed a strong co-expression (80.45%) among the down regulated genes (Figure 6B). Protein localization to synapse, regulation of neurotransmitter receptors, glutamatergic synapse, and neuron projection were among the most significant pathways enriched in the down-regulated genes shared between the *MAPT* mutant iPSC-neurons and PSP brains (Supplemental Table 13). The up-regulated genes (n=33) were enriched in pathways involved in negative regulation of neuronal projection and trans-synaptic signaling (Figure 6C). Together, these analyses identified a set of gene signatures captured in stem cell models of *MAPT* mutations that also change in human brains with sporadic tauopathy pathology.

**Figure 6.**
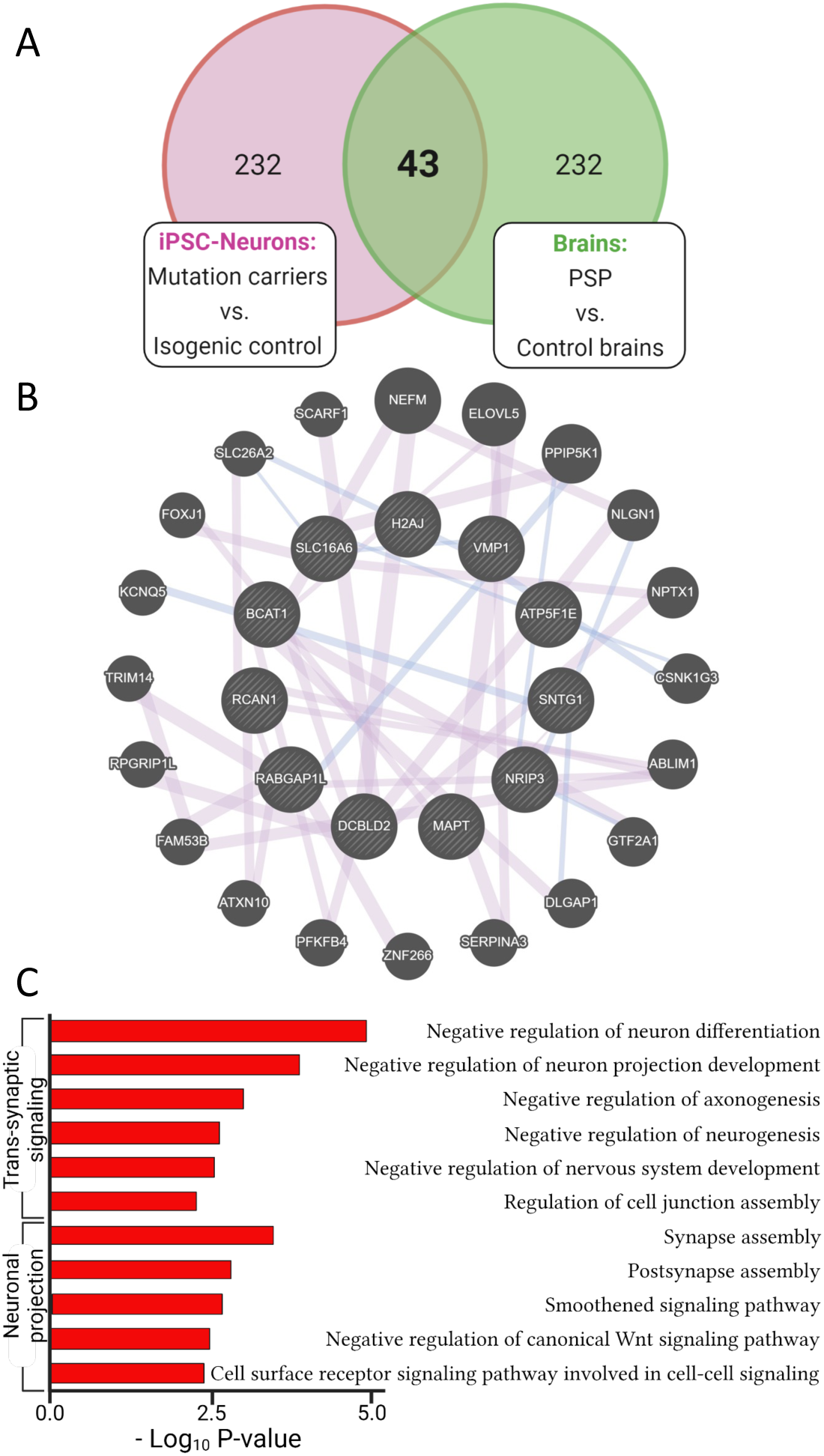
Genes changed in iPSC-neurons from *MAPT* mutations are also altered in sporadic tauopathies. (A) Venn diagram representing the 43 differentially expressed genes (33 up- and 10 down-regulated genes) shared between cell lines expressing *MAPT* IVS10+16, p.P301L and p.R406W mutations (275 genes; p ≤ 0.05) and progressive supranuclear palsy (PSP) brains. (B) Relatedness of the 10 down-regulated genes overlapped between cell lines carrying *MAPT* IVS10+16, p.P301L and p.R406W mutations and PSP brains (p ≤ 0.05), including the *MAPT* gene. The sizes of the gene nodes are proportional to the degrees to which the genes are related. While query genes are presented as striped dark grey balls, other selected genes subjected to interaction are presented as dark grey balls without stripes. Purple links represent genes co-expressed (80.45%), and blue lines show co-localized genes (19.55%). (C) Bar graph showing the most significant pathways enriched among the 33 up-regulated genes (red bars) overlapped between the three mutations and the PSP brains.

**Figure 7.**
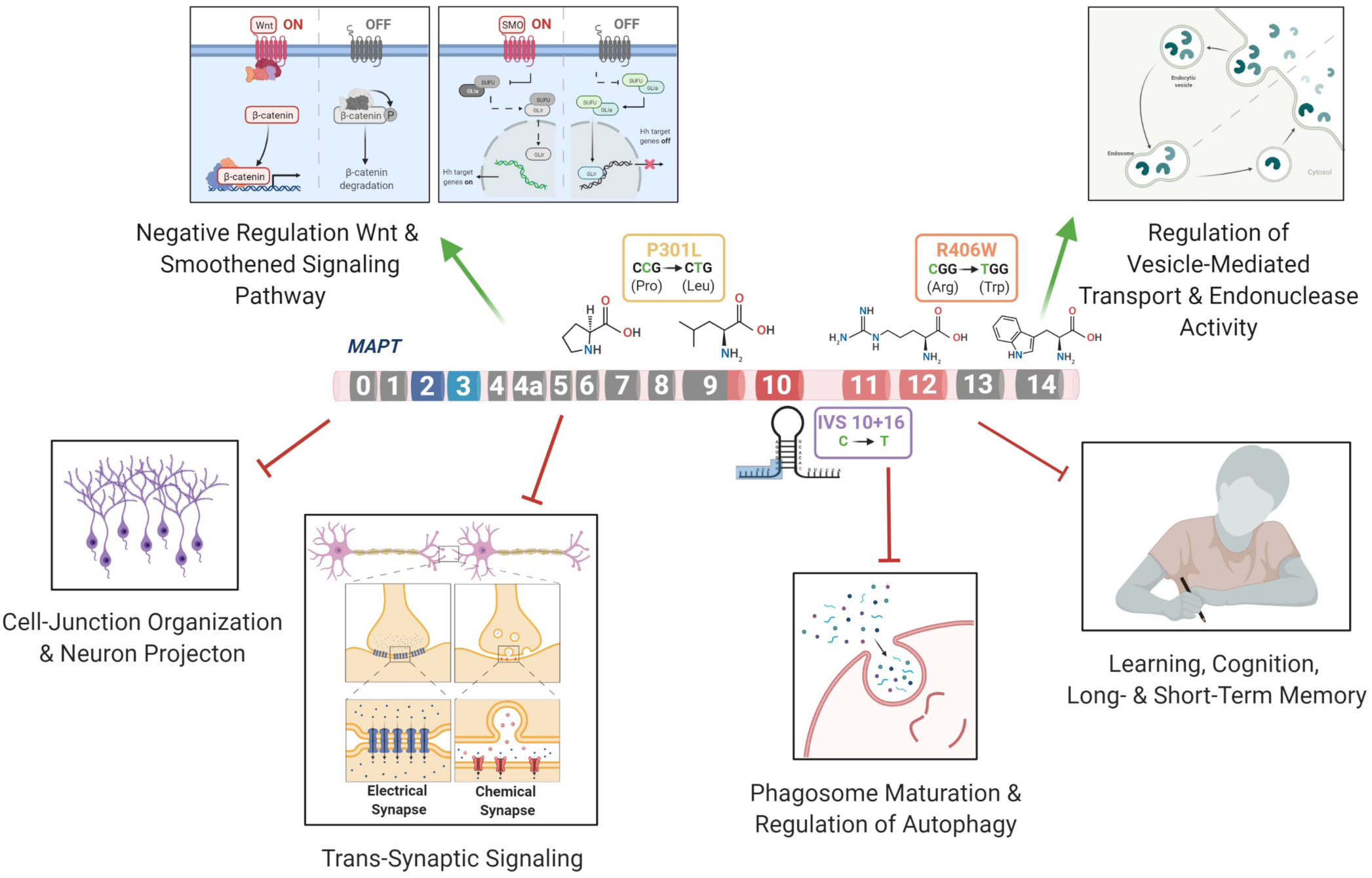
*MAPT* Mutations Lead to Changes in the Regulation of Pathways related to FTLD- Tau.

## Discussion

The goal of this study was to identify commonly perturbed genes and pathways downstream of the *MAPT* mutation and to define a core set of genes that drive disease pathogenesis in FTLD-Tau. *MAPT* mutations result in a range of clinical and neuropathological phenotypes (Moore *et al*., 2020; Whitwell *et al*., 2009; Ghetti *et al*., 2015). Here, we aimed to study three major classes of *MAPT* mutations that represent: splicing mutation (*MAPT* IVS10+16), 4R-expressing point mutation (*MAPT* p.P301L) and point mutation expressed in all isoforms (*MAPT* p.R406W). Our results suggest that these three mutations lead to a common series of events, causing the dysregulation of genes associated with pathways involved in synaptic function, and endolysosomal pathways, and neuronal development. Several of the differentially expressed genes were also altered in a mouse model of tauopathy, suggesting that these genes are relevant to disease pathogenesis and accumulation of NFTs. A subset of genes were also found to be dysregulated in human brains from progressive supranuclear palsy (PSP) patients, illustrating that the signatures we identified in iPSC-neurons are relevant to the human disease. Thus, this study demonstrates that iPSC-derived neurons capture molecular processes that occur in both mice and human brains can be used to model neurodegenerative diseases such as FTLD-tau.

The three mutations located in distinct regions of the *MAPT* gene produced a common molecular signature of 275 genes that was enriched for pathways involved in trans-synaptic signaling, neuronal projection and endolysosomal regulation. Among the dysregulated pathways associated with the 159 up-regulated genes, several pathways were related to functional changes described in FTLD. Wnt signaling has been previously shown to be associated with the *MAPT* IVS10+16 mutation (Harrison-Uy and Pleasure, 2012) and linked to several neurodegenerative disorders such as AD and FTD (Riise *et al*., 2015; Rosen *et al*., 2011; Korade and Mirnics, 2011; Verheyen *et al*., 2018; Bottero *et al*., 2021). In addition, Notch signaling has been shown to be related to the microtubule stability within neurons (Bonini *et al*., 2013). The Ras signaling regulates basic cellular processes in the construction of neuronal networks, including neurogenesis, vesicular trafficking, or synaptic plasticity (Johnson and Chen, 2012; Qu *et al*., 2019). The synapse assembly has been associated with Alzheimer’s-type dementia (Clare *et al*., 2010). The 116 commonly down-regulated genes were enriched for synaptic pathways. Anterograde trans-synaptic signaling; regulation of synaptic; and long-term synaptic potentiation have all been implicated in tauopathies (Liou *et al*., 2019; Gómez-Palacio-Schjetnan and Escobar, 2013; Purves, Augustine and Fitzpatrick, 2001). Additionally, several genes were enriched for pathways related to the loss of learning or memory and cognition (Rabinovici and Miller, 2010; Bott *et al*., 2014). Finally, we found that genes associated with dysregulation of the vesicle transport along microtubule pathway; phagosome maturation and autophagy were significantly reduced. This finding is consistent with a recent study of *MAPT* p.V337M-expressing cerebral organoids, where a loss of protein homeostasis was reported (Bowles *et al*., 2021). Defects in endolysosomal pathways have been implicated in reduced clearance of protein and cell debris, which may contribute to neurodegeneration (Liu, Rizzo and Puthanveettil, 2012; Gunawardena, Anderson and White, 2014; Etchegaray *et al*., 2016; Viegas, Estronca and Vieira, 2012; Kulkarni and Maday, 2018; Xu *et al*., 2021). Thus, here, we identify a series of pathways that are common across different *MAPT* mutation types, suggesting that stem cell models capture defects observed in FTLD-tau patients.

We have shown that several of the commonly differentially expressed genes from the mutant iPSC-neurons were also altered at disease end-stage in the Tau-P301L mouse model of tauopathy: *CELSR1, CHRDL1, CALB1, PLK2, PRICKLE2,* and *ST8SIA3.* FDA approved drugs were identified for two targets, *PLK2* and *CALB1*: Ethosuximide, Oxcarbazepine, and Levodopa (Chen *et al*., 2015; Tambasco, Romoli and Calabresi, 2018; Morana *et al*., 2020). *CELSR1* (Cadherin EGF LAG seven-pass G-type receptor 1) encodes a receptor protein involved in cell adhesion and receptor-ligand interactions (Hadjantonakis *et al*., 1997). *CELSR1* has been related to neurodevelopment and maintenance of the nervous system (Boutin, Goffinet and Tissir, 2012), and mutations in this gene has been associated with neural tube defects (Robinson *et al*., 2012) and AD risk (Patel *et al*., 2019). *CHRDL1* (Chordin-like 1) plays an important role in CNS development, learning, and promoting synaptic plasticity (Sun *et al*., 2007; Gao *et al*., 2013; Webb *et al*., 2012; Blanco-Suarez *et al*., 2018). *PLK2* (Polo like kinase 2) is associated with synaptic plasticity and prevention of cell death in neurodegenerative diseases (Kauselmann, 1999; Seeburg, Pak and Sheng, 2005; Seeburg *et al*., 2008; Li *et al*., 2014; Weston *et al*., 2021). Modulating the activity of the *PLK2* gene has been proposed as a therapeutic strategy for the treatment of Parkinson’s disease (Oueslati *et al*., 2013). *PRICKLE2* (Prickle planar cell polarity protein 2) is an important cytoplasmic regulator of Wnt/PCP signaling (Vandervorst *et al*., 2019; Katoh, 2005; Barrow, 2006). Dysregulation of *PRICKLE2* enhances the amyloid β (Aβ) plaque pathology and synaptic dysfunction in mice (Fujimura and Hatano, 2012; Tao *et al*., 2011). *PRICKLE2* has been suggested to be a potential candidate for the diagnosis and treatment of AD (Sun *et al*., 2020). (iv) *ST8SIA3* (alpha-N-acetyl-neuraminide alpha-2,8-sialyltransferase 3) is involved in neurite growth, cell migration, and synaptic plasticity (Lee *et al*., 1998; Eckhardt *et al*., 2000; Goodman, Davis and Zito, 1997; Lin *et al*., 2019), and plays an important role in the development of Huntington’s disease, schizophrenia, and Parkinson’s disease (Moll, Shaw and Cooper-Knock, 2020; Belarbi *et al*., 2020). A strong physical interaction was observed between *CELSR1, CHRDL1, CALB1, PLK2, PRICKLE2, ST8SIA3, MAPT* and other 20 additional genes. The glutamatergic synapse pathway was enriched among these genes. Impairments in glutamatergic circuits predispose GABAergic neurons to dysfunction (Benussi *et al*., 2019; Bowie, 2008; Murley *et al*., 2020; Ferrer, 1999; Hughes *et al*., 2018). Dysregulation of the glutamatergic system has been described in *MAPT* p.V337M (Borroni *et al*., 2017; Borroni *et al*., 2018; Bowles *et al*., 2021) and p.R406W (Jiang *et al*., 2018). Here, our observations link the dysfunction of the glutamatergic synapse pathway to the *MAPT* mutations IVS10+16, p.P301L, and p.R406W.

*CALB1* was found to be down regulated across the three *MAPT* mutations and reduced at disease end-stage in Tau-P301L mice. The *CALB1* gene (Calbindin 1) regulates the calcium homeostasis in neurons (Noble *et al*., 2018), which plays a crucial role in neuronal development and memory performance (Jung *et al*., 2020; Sun *et al*., 2005; Kook *et al*., 2014; Goffigan-Holmes *et al*., 2018; Soontornniyomkij *et al*., 2012). Given the important role of *CALB1* in neuronal calcium homeostasis, we measured calcium levels in the mutant and isogenic control neurons. We found that calcium levels were significantly reduced in *MAPT* mutant neurons compared with isogenic controls, supporting a dysregulation in calcium homeostasis. Calcium homeostasis is critical for the health and function of neurons and dysregulation of calcium leads to altered synaptic function, endolysosomal function, and neuronal development (Gleichmann and Mattson, 2011). This is in line with recent observations in genetically engineered iPSC expressing the *MAPT* IVS10+16 mutation, which showed disturbed intracellular calcium dynamics along with impaired neuronal activity (Kopach *et al*.; Britti *et al*.). Taken together, we were able to re-capitulate some of these disease phenotypes in the patient-derived neurons harboring the MAPT IVS10+16 mutation.

A subset of the commonly differentially expressed genes from the mutant iPSC-neurons were found to be altered in human brains with the primary tauopathy, PSP (Allen *et al*., 2016). Among the up-regulated genes, we observed an enrichment in pathways involving a negative regulation of: (i) nervous system development; (ii) neurogenesis; (iii) neuron differentiation; (iv) neuron projection development; and (v) the canonical Wnt signaling pathway. Dysregulation of these pathways have been linked to FTLD patients (Yousef *et al*., 2017; Rabinovici and Miller, 2010; Mann and Snowden, 2017; Sobue, Ishigaki and Watanabe, 2018; Bottero *et al*., 2021). The down-regulated genes *VMP1, RABGAP1L* were enriched for the protein transport pathway, which affects the nucleocytoplasmic shuttling inside the neurons, and it has been associated with FTLD (Prpar Mihevc *et al*., 2017; Karamysheva, Tikhonova and Karamyshev, 2019). Genes found in our network analyses were associated with pathways such as: (i) microtubule-based transport; (ii) neuron to neuron synapse; (iii) regulation of canonical Wnt signaling; and (iv) regulation of neurotransmitter receptor activity. All these pathways have been associated with abnormalities in the nervous system development (Rabinovici and Miller, 2010; Sobue, Ishigaki and Watanabe, 2018) and loss in intracellular transport and signaling (Liu, Rizzo and Puthanveettil, 2012; Murley and Rowe, 2018; Benussi *et al*., 2019). Thus, the stem cell model revealed several genes and pathways that are also altered in primary tauopathy patients.

Our results demonstrate the potential of the iPSC technology to investigate disease mechanisms related to FTD neurodegeneration. A major challenge related to the iPSC technology and neuronal derivatives (iPSC-neurons) when modeling adult-onset neurodegenerative disease concerns capturing the age-related phenotypes (Steg *et al*., 2021). While iPSC-neurons remain relatively immature and do not express the full complement of Tau isoforms (Sato *et al*., 2018; Sposito *et al*., 2015), we are able to capture molecular signatures that change during disease course in animal models of tauopathy and patient brains. Thus, there remains tremendous value in a combinatorial multiple model systems approach to identify key pathways that are affected early and remain relevant throughout the disease course.

In conclusion, our approach provides a tractable system to identify genes altered directly by the *MAPT*-IVS10+16, p.P301L, and p.R406W mutations, which are relevant to tauopathies and that point to new therapeutic targets. The stem cell lines used in this research allowed for the identification of molecular drivers of disease, which could serve as a platform to identify new targets for drug development. Our iPSC-based cellular models have discovered a common gene signature which is enriched in dysregulated pathways involving synaptic connections, lysosome transport and neuronal development, and mechanisms that have been previously described to be altered in PSP brains, and mouse models of tauopathy.

## Data Availability

All data produced in the present study are available upon reasonable request to the authors

## Acknowledgements

We would like to thank the research subjects and their families who generously participated in this study. We thank Dr. Aimee Kao who generously provided the fibroblasts used to generate the GIH36 iPSC line (supported by the Rainwater Charitable Organization). This work was supported by access to equipment made possible by the Hope Center for Neurological Disorders, the Neurogenomics and Informatics Center, and the Departments of Neurology and Psychiatry at Washington University School of Medicine. OH is an Archer Foundation Research Scientist. Confocal images were generated on a Zeiss LSM 880 Airyscan Confocal Microscope which was purchased with support from the Office of Research Infrastructure Programs (ORIP), a part of the NIH Office of the Director under grant OD021629. Funding provided by the National Institutes of Health (P30 AG066444, R01 AG056293, R56 NS110890, RF1 NS110890, U54 NS123985, R01 AG062359, R01 AG057777), Hope Center for Neurological Disorders (CMK), Rainwater Charitable Organization (CMK), Farrell Family Fund for Alzheimer’s Disease (CMK), and UL1TR002345. The recruitment and clinical characterization of research participants at Washington University were supported by NIH P30AG066444 (JCM), P01AG03991 (JCM), and P01AG026276 (JCM). Diagrams were generated used BioRender.com.

## Supplemental Figures

**Supplemental Figure 1:**
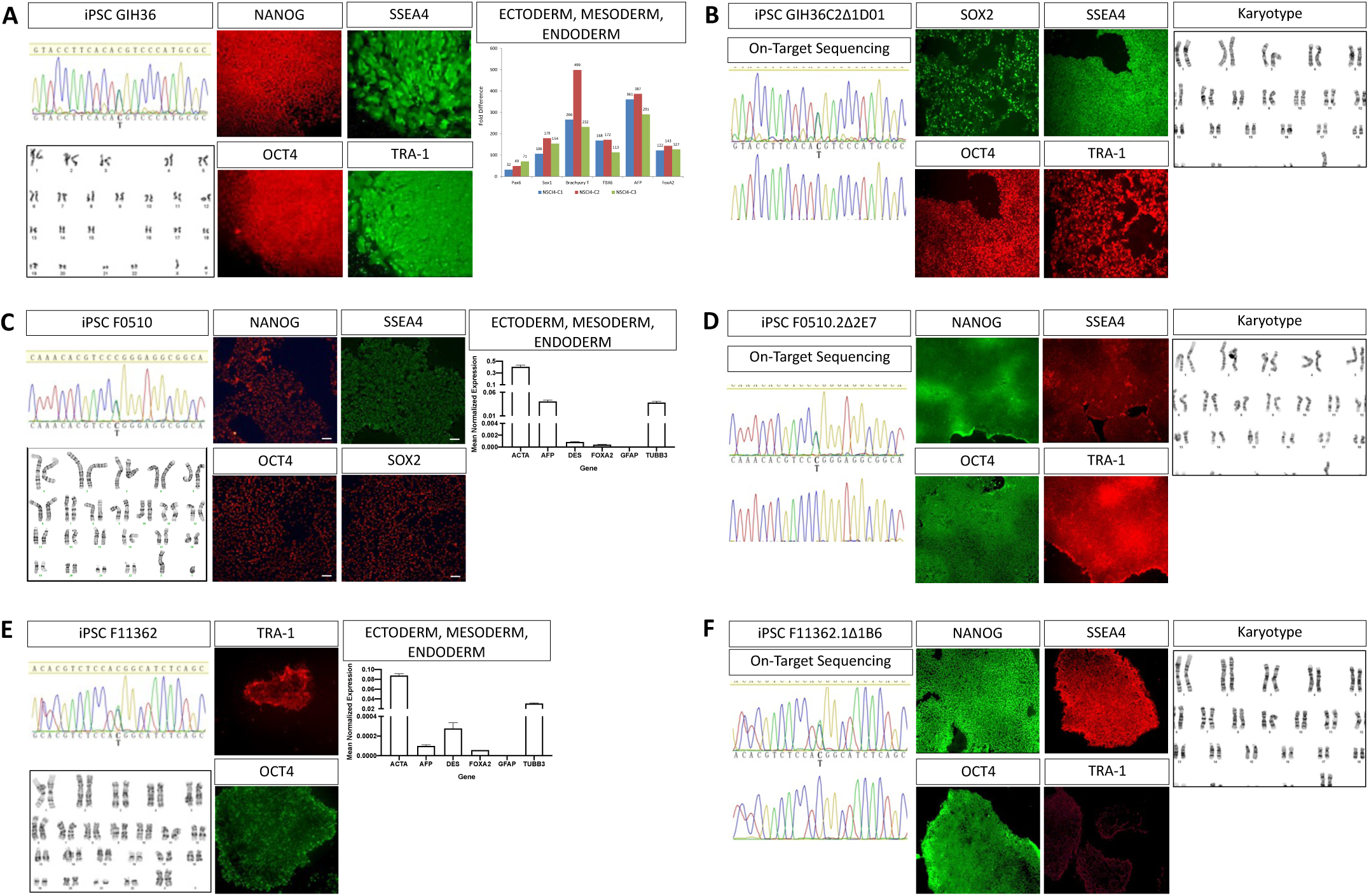
Characterization of iPSC from *MAPT* IVS10+16, p.P301L, and p.R406W and isogenic controls. Sanger sequencing, karyotyping, and immunocytochemistry for pluripotency for patient lines and isogenic controls. For patient lines, qPCR for spontaneous differentiation of iPSC into ectoderm, mesoderm, and endoderm cell-types. (A) GIH36.2. (B) GIH36C2Δ1D01. (C) F5010.2. (D) F0510.2Δ2’H1. (E) F11362.1. (F) F11362.1Δ1B6.

**Supplemental Figure 2.**
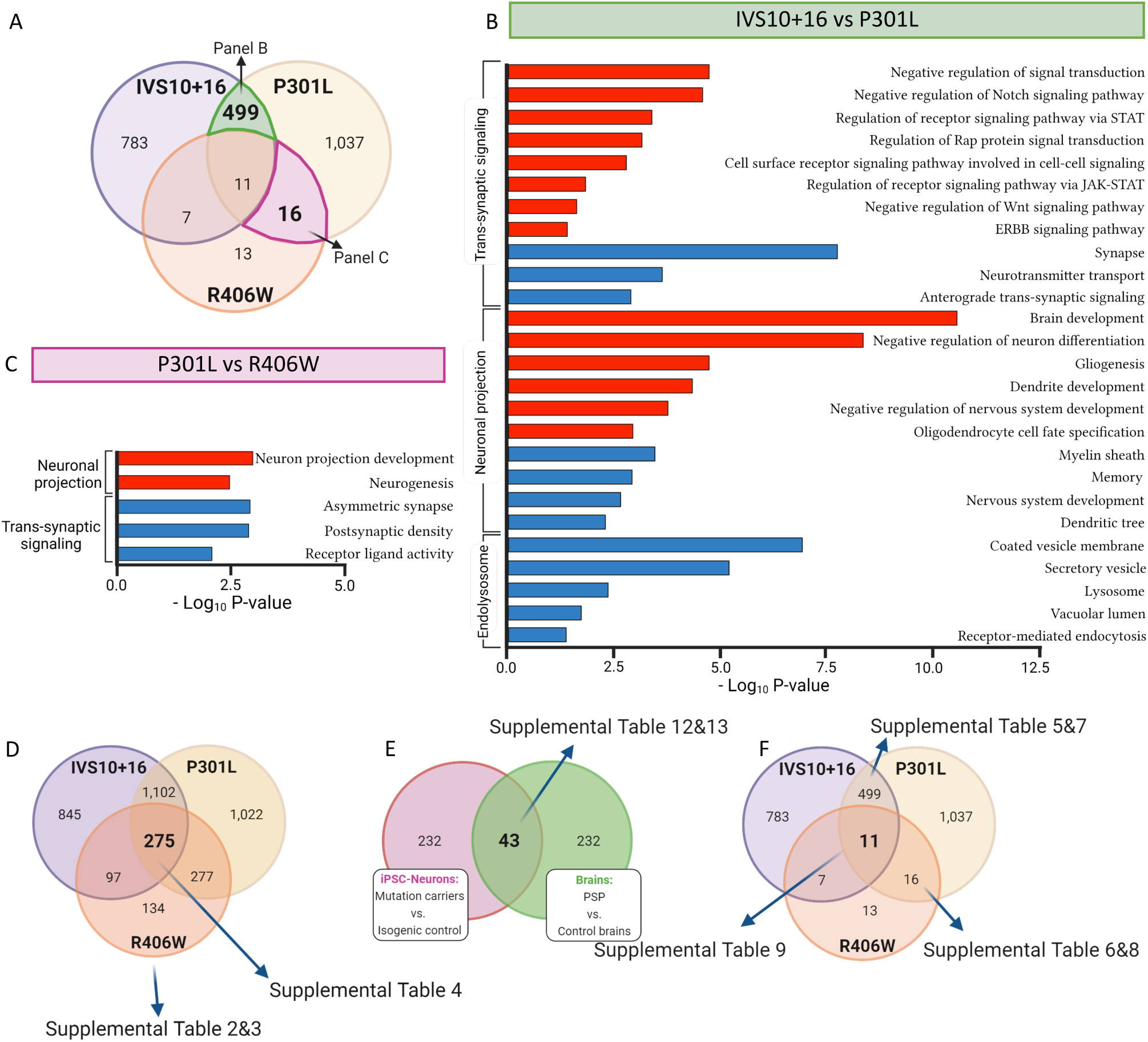
Common gene signatures among exon 10 (IVS10+16/P301L) or point mutations (R406W/P301L). (A) Venn diagram overlapping the differentially expressed genes obtained from iPSC-neurons carrying *MAPT-*IVS10+16, p.P301L and p.R406W mutations (compared to isogenic controls). Genes were previously selected using adjusted BY-FDR ≤ 0.05. The green highlighted sector shows the 499 overlapped genes overlapped between IVS10+16 (*MAPT* intron 10) vs P301L (*MAPT* exon 10). The purple highlighted sector presents the 16 overlapped genes between P301L vs R406W (*MAPT* exon 13). (B) Bar graph shows pathways enriched with the 235 up- and 264 down-regulated genes overlapped between IVS 10+16 and P301L. Up-regulated genes (red bars) were enriched in pathways related to negative regulation of trans-synaptic signaling and negative regulation of nervous system development. Down-regulated genes (blue bars) were enriched in pathways associated to trans-synaptic signaling, nervous system development and endolysosomal function. (C) Bar graph showing pathways enriched with the 2 up- and 14 down-regulated genes overlapped between P301L and R406W. While the up-regulated genes (red bars) were associated to pathways related to neuron projection, the down regulated genes (blue bars) were associated with trans-synaptic signaling. (D-F) Diagrams illustrating the gene comparisons included in Supplemental Tables 2, 3 and 4 (D); Supplemental Tables 12 and 13 (E); and Supplemental Tables 9 and 5-8 (F).

**Supplemental Figure 3.**
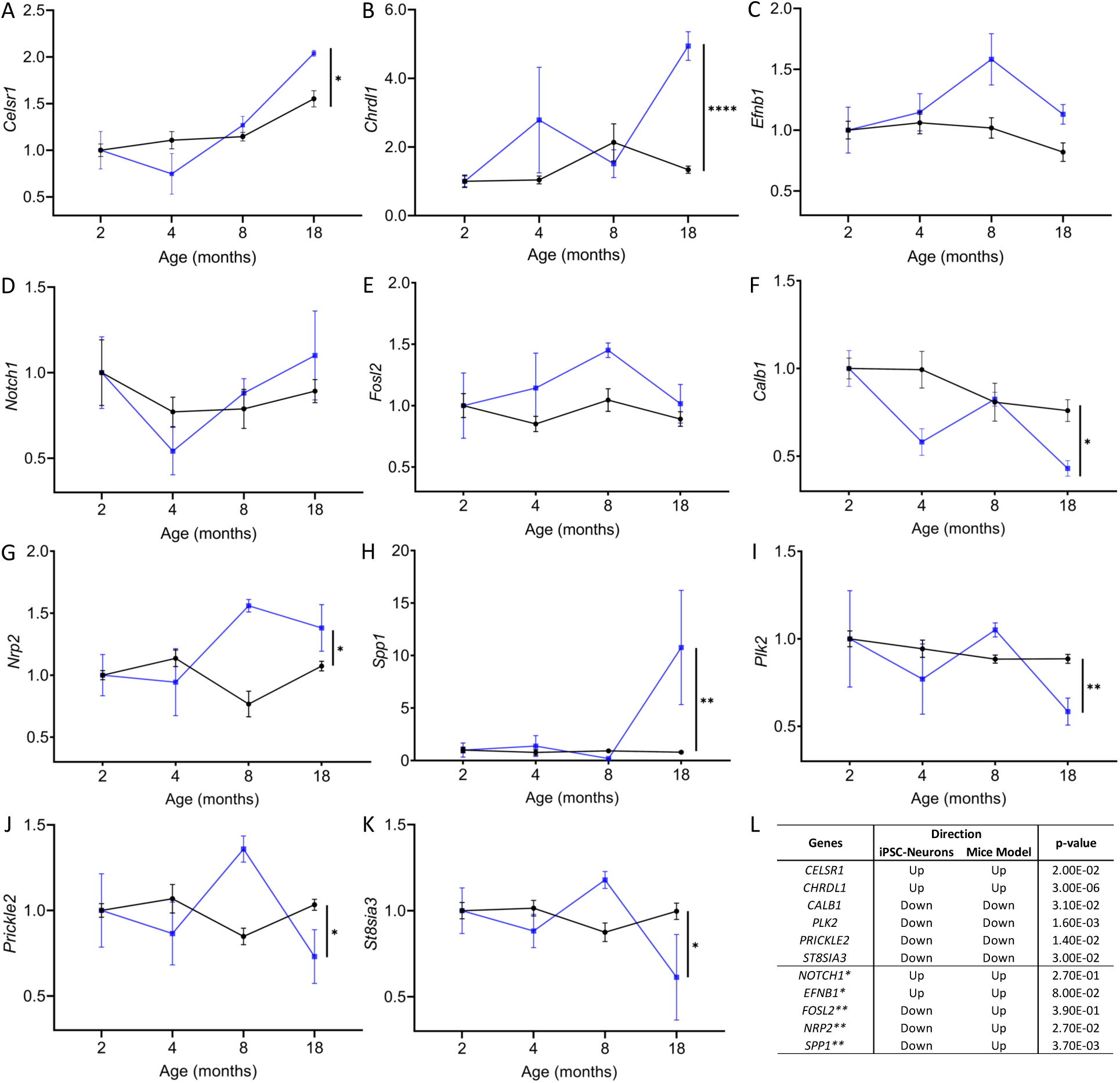
Gene expression in the Tau-P301L transgenic mouse model of tauopathy. (A – K) The 11 genes (BY-FDR ≤ 0.05) observed in neurons carrying *MAPT-*IVS10+16, p.P301L and p.R406W mutations were examined in a database containing gene expression from transgenic Tau-P301L (black circles) and non-transgenic control mice (blue squares). Graphs show normalized gene expressions related to data obtained in mice that were two months old. Statistical comparisons were done on values obtained from 18 months old mice when Tau aggregation is most robust. *p ≤ 0.05; **p ≤ 0.005; ****p ≤ 0.00005. (L) Summary statistics of analyses in mice. *t-test p > 0.05; ** Genes exhibiting different directions of effect between iPSC-neurons and mice.

